# Comparative study of immunogenicity and safety of Gam-COVID-Vac and Sinopharm BBIBP-CorV vaccines in Belarus

**DOI:** 10.1101/2022.02.05.22270499

**Authors:** Igor Stoma, Katsiaryna Korsak, Evgenii Voropaev, Olga Osipkina, Aleksey Kovalev

## Abstract

**Introduction:** Lack of comparative studies on efficiency of a broad range of COVID19 vaccines leads to lower levels of adoption and subsequent lower total immunity in several regions, including Republic of Belarus. This clinical study captures and transparently demonstrates varying immunogenic responses to Sputnik V and Sinopharm vaccines.

**Aim of this study was:** to compare the immunogenicity and reactogenicity of Sputnik V (Gam-COVID-Vac), RF and Sinopharm (BBIBP-CorV), PRC vaccines in vaccinated individuals.

**Materials and Methods:** A total of 60 adults participated the study. The immune response after vaccination was assessed using enzyme immunoassay. IgG levels measured in all participants at three time points: before the vaccination, 42 days after the first vaccine dose, and 6 months after the first vaccine dose. The results of the SARS-CoV-2 antibody test is quantified according to the WHO First International Standard (NIBSC code:20/136) and expressed in international units (BAU/ml).

**Results:** The study participants were divided into two groups, where 30 people (50%) were vaccinated with Sputnik V (Gam-COVID-Vac), and 30 people were vaccinated with Sinopharm (BBIBP-CorV), with no gender differences in the groups. The IgG levels at 42 days after the first vaccine dose were: Sputnik V (Gam-COVID-Vac)_(42 days)_: Me=650.4 (642.2-669.4); Sinopharm (BBIBP-CorV)_(42 days)_: Me=376.5 (290.9-526.4); p<0,001). The IgG levels at 6 months after the first vaccine dose were: Sputnik V (Gam-COVID-Vac)_(6 months)_ Me=608.7 (574.6-647.1); Sinopharm (BBIBP-CorV)_(6 months)_: Me=106.3 (78.21-332.4); p<0,001). Reactions after vaccination appeared in 27 vaccinated individuals (45%).

**Conclusion:** The study showed that Sputnik V (Gam-COVID-Vac) vaccine was more immunogenic than Sinopharm (BBIBP-CorV) vaccine. IgG levels in vaccinated individuals who previously recovered from SARS-CoV-2 infection (“hybrid immunity”) were higher than in SARS-CoV-2 naïve individuals. Reactions after vaccines administration were mild to moderate.

## Introduction

The pandemic caused by SARS-CoV-2 has been ongoing for two years. Due to the lack of etiotropic therapy for COVID-19 [1], increased infectivity [2] and the acquisition of vaccine-resistant mutations of SARS-CoV-2 [3], vaccination is the only effective means of controlling the epidemic. It has the potential to curb the spread of the virus [4], as well as to reduce the consequences of infection and the burden on health care facilities [5]. Effective pandemic control requires a high vaccination rate [6].

To date, 33 vaccines against SARS-CoV-2 have been approved for mass administration worldwide [7]. They differ in their manufacturing technology, their mechanism of action and efficacy.

In the Republic of Belarus, currently two vaccines are available to the population: Sputnik V (Gam-COVID-Vac), manufactured in the Russian Federation and Sinopharm (BBIBP-CorV), manufactured in China. As of 01.02.22, 45.0% of population were vaccinated with two doses, 57.4% received one dose of vaccine and 2.9% were revaccinated [8]. There is a lack of studies that compare the properties of vaccines (immunological efficacy, postvaccination reaction rates) Sputnik V (Gam-COVID-Vac), RF and Sinopharm (BBIBP-CorV), PRC between each other in the international database of medical publications. The lack of comparative information about available vaccines is one of the reasons why the required vaccination coverage has not yet been achieved in the Republic of Belarus

### Aim of the study

To compare the immunogenicity and reactogenicity of Sputnik V (Gam-COVID-Vac), RF and Sinopharm (BBIBP-CorV), PRC vaccines among vaccinated individuals.

## Materials and methods

This prospective clinical study was carried out at Gomel State Medical University, Gomel, Belarus from October 2020 to December 2021. Evaluation of the immune response to vaccination was performed by enzyme immunoassay using the Sunrise Tecan microplate photometer (Austria). Blood sampling was performed 3 times: immediately before the first dose of vaccine, on day 42 and 6 months after the first dose of vaccine. All study participants were informed about the study design and upcoming procedures, all participants obtained informed written consent to participate in the study. The study received an approval from the institutional Ethics committee of Gomel state medical university. Vaccination was performed in a strict adherence to the vaccines instruction (product medical information) and in accordance with principles of GDP (good distribution practice), with 2 doses of vaccines administered to all of the participants.

A reagent kit manufactured by Vector-Best (Russian Federation) and designed for enzyme immunoassay for quantitative detection of SARS-CoV-2 class G immunoglobulins, SARS-CoV-2-IgG quantitative ELISA-BEST, was used for immunoassay. According to the manufacturer’s information letter, this kit is suitable for the quantitative detection of specific IgG in SARS-CoV-2 infected and re-vaccinated patients as well as for the evaluation of post-vaccination immune response during immunization with vaccines based on different technologies (vector, mRNA, inactivated whole-virion). The “SARS-CoV-2-IgG quantitative ELISA-BEST” reagent kit design uses recombinant full-length trimerized S glycoprotein (Spike) of the SARS-CoV-2 virus derived from a eukaryotic expression system. The protein molecule+ consists of two subunits, S1 containing the RBD domain and S2. The reagent kit “SARS-CoV-2-IgG quantitative ELISA-BEST” detects the pool of immunoglobulin class G synthesized to all antigenic determinants of protein S including the RBD-domain.

The quantification of the SARS-CoV-2 antibody assay is based on the WHO First International Standard (NIBSC code:20/136) and expressed in international units (BAU/ml).

According to the manufacturer’s instructions, the diagnostic sensitivity for the detection of IgG to SARS-CoV-2 is 100% (range 95.7%-100%, 95% confidence interval). The diagnostic specificity is 100% (range 98.5%-100%, 95% confidence interval). The information letter accompanying the reagent kit “SARS-CoV-2-IgG Quantitative ELISA-BEST”, No. RZN 2021/14458 reports that virus neutralizing activity with a neutralization titer of 1/160 or higher is observed in all samples with a specific IgG concentration of 150 BAU/ml or higher (ELISA titer ≥1/600) (95% CI: 83.16 - 100%) and only 50% of samples with a specific IgG concentration of 80-149 BAU/ml (ELISA titer ≥1/400 - 1/800) (95% CI: 32.43 - 67.57%), a specific IgG level of less than 10 BAU/ml should be considered as a negative result of the quantitative assay.

On every visit participants filled out a questionnaire including data on the course of chronic diseases, history of COVID-19, and adverse reactions (if any). In case of any adverse reactions due to vaccination the principal investigator was informed immediately.

Statistical processing of the results was performed using the R statistical programming environment (graphs and statistical criterion calculations were obtained using the basic R package and the ggpubr package). Normality of the distribution was assessed using the Shapiro-Wilk test. Quantitative comparisons of linked samples (change in IgG levels over time in groups with different vaccines (Sputnik V (Gam-COVID-Vac), RF and Sinopharm (BBIBP-CorV), PRC) were performed using the Friedman test, followed by pairwise comparison of groups by the Wilcoxon test with Bonferroni multiple comparison adjustment. Mann-Whitney test was used for unrelated samples (comparison of the dynamics of IgG levels in the group in which participants were vaccinated with Sputnik V (Gam-COVID-Vac), RF with the group vaccinated with Sinopharm (BBIBP-CorV), China). A qualitative comparison of the groups (achievement of one of the IgG values: up to 150 BAU/ml, 150-500 BAU/ml, >500 BAU/ml after administration of Sputnik V vaccine (Gam-COVID-Vac), RF and Sinopharm (BBIBP-CorV) vaccine, China; incidence of major postvaccination reactions (temperature, injection site soreness) among participants vaccinated with Sputnik V vaccine (Gam-COVID-Vac), RF and Sinopharm vaccine (BBIBP-CorV), China) was estimated using the Pearson χ2 test; Fisher’s Exact Test was used in cases where expected values in the cells of the contingency table were less than 5. The significance level was set at 0.05.

## Results

There were in total 60 participants in this study. Males accounted for 30% (18 participants) and females accounted for 70% of the participants (42 participants). Study participants were divided into two groups, where 30 people (50%) were vaccinated with Sputnik V (Gam-COVID-Vac), RF (11 men (36.67%), 19 women (63.33%)), 30 people were vaccinated with Sinopharm (BBIBP-CorV) vaccine, PRC (7 men (23.33%), 23 women (76.67%)). According to the questionnaire, 43 people (71.67%) reported having no chronic diseases. The chronic diseases indicated can be grouped as follows: 1) cardiovascular diseases were 35.29% (6 individuals), 2) gastrointestinal diseases were 35.29% (6 individuals), 3) musculoskeletal diseases were 17.65% (3 individuals), 4) respiratory diseases were 11.76% (2 individuals), 5) endocrine diseases were 5.88% (1 individual), 6) female reproductive system diseases were 5.88% (1 individual). Participants in the study reported more than one chronic disease. When comparing the two groups of participants on the basis of chronic diseases, no significant difference was found (p=1.0). The characteristics of the study participants are presented in Table 1.

**Table 1:**
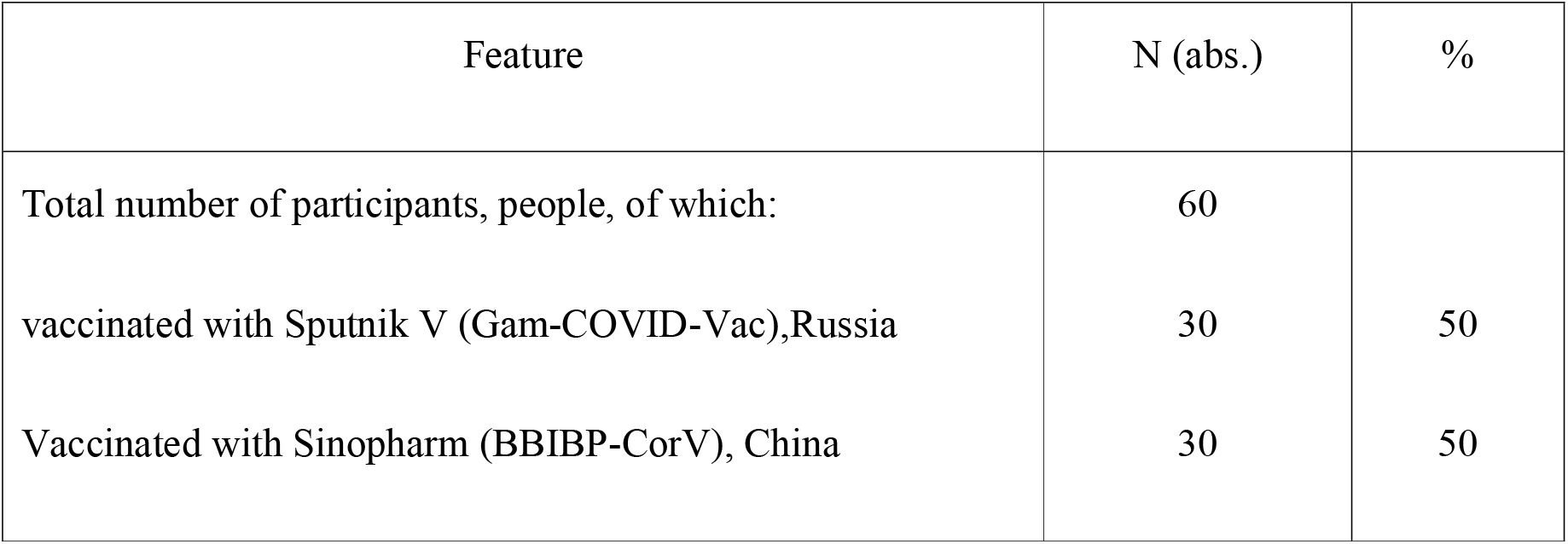
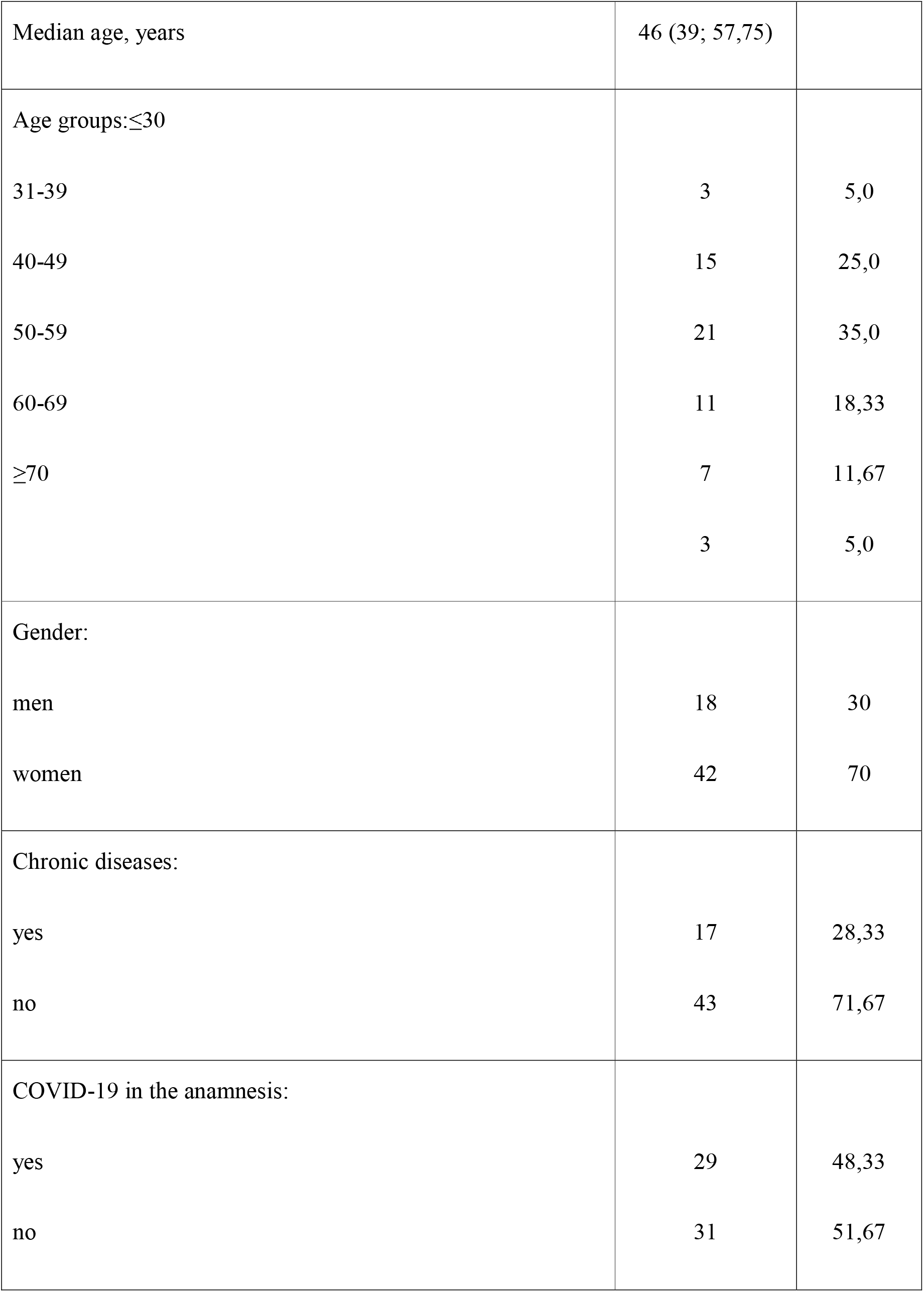
Characteristics of study participants.

The main surrogate marker for a vaccine efficacy is its immunogenicity. In this study, IgG levels to SARS-CoV-2 S-protein were tested in all participants at three stages: before vaccination, 42 days after the first dose of vaccine and 6 months after the first dose of vaccine. The unit of measure for IgG levels is BAU/ml. (Note: BAU-binding antibody units). The study data are shown in Figure 1, Figure 2 and Figure 3.

**Fig. 1.**
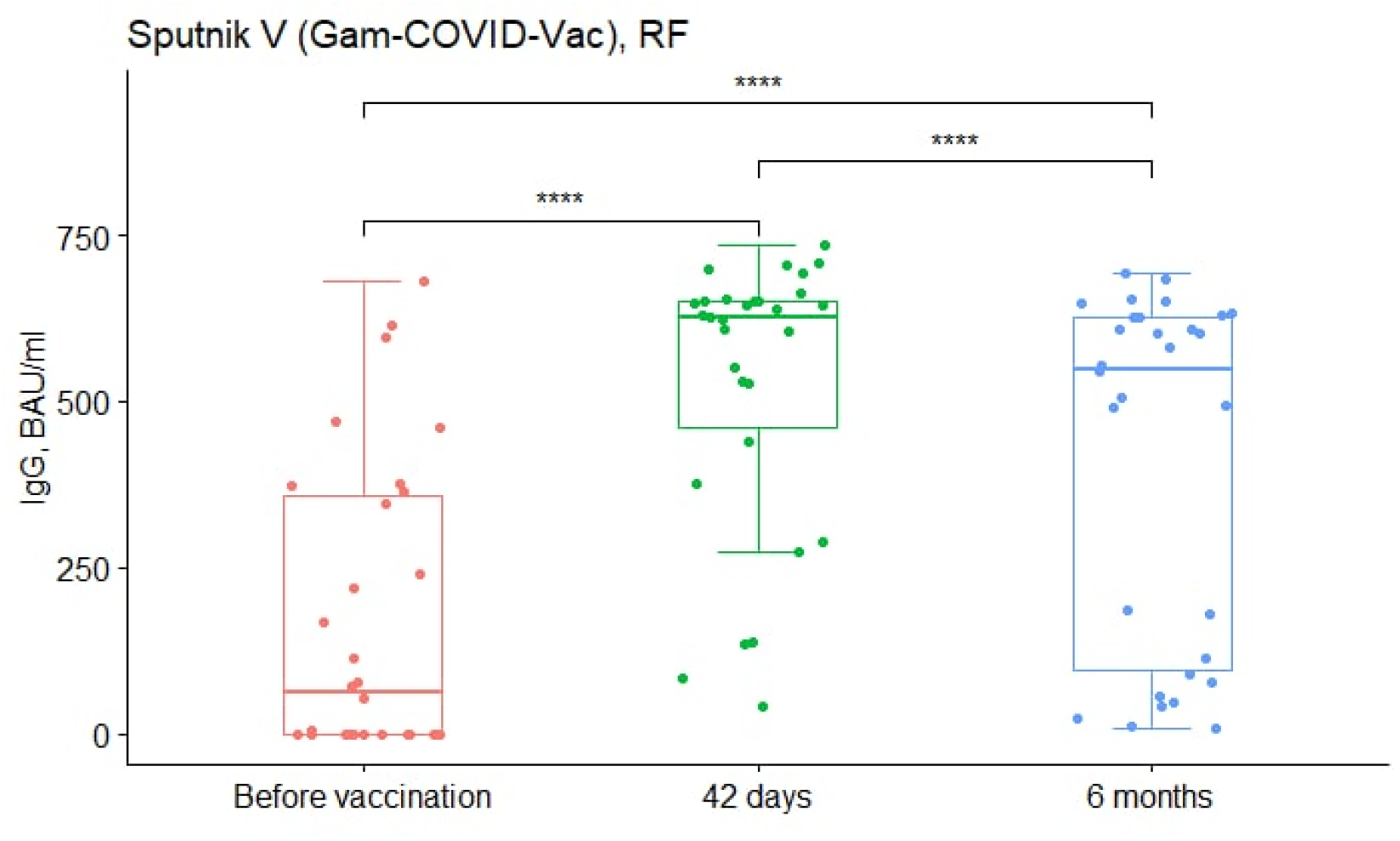
Dynamics of Ig G production in response to Vaccine Sputnik V (Gam-COVID-Vac). (“before vaccination” - Ig G value (BAU/ml) before vaccination; “42 days” - Ig G value (BAU/ml) 42 days after the first dose; “6 months” - Ig G value (BAU/ml) 6 months after the first vaccine dose).

**Fig. 2.**
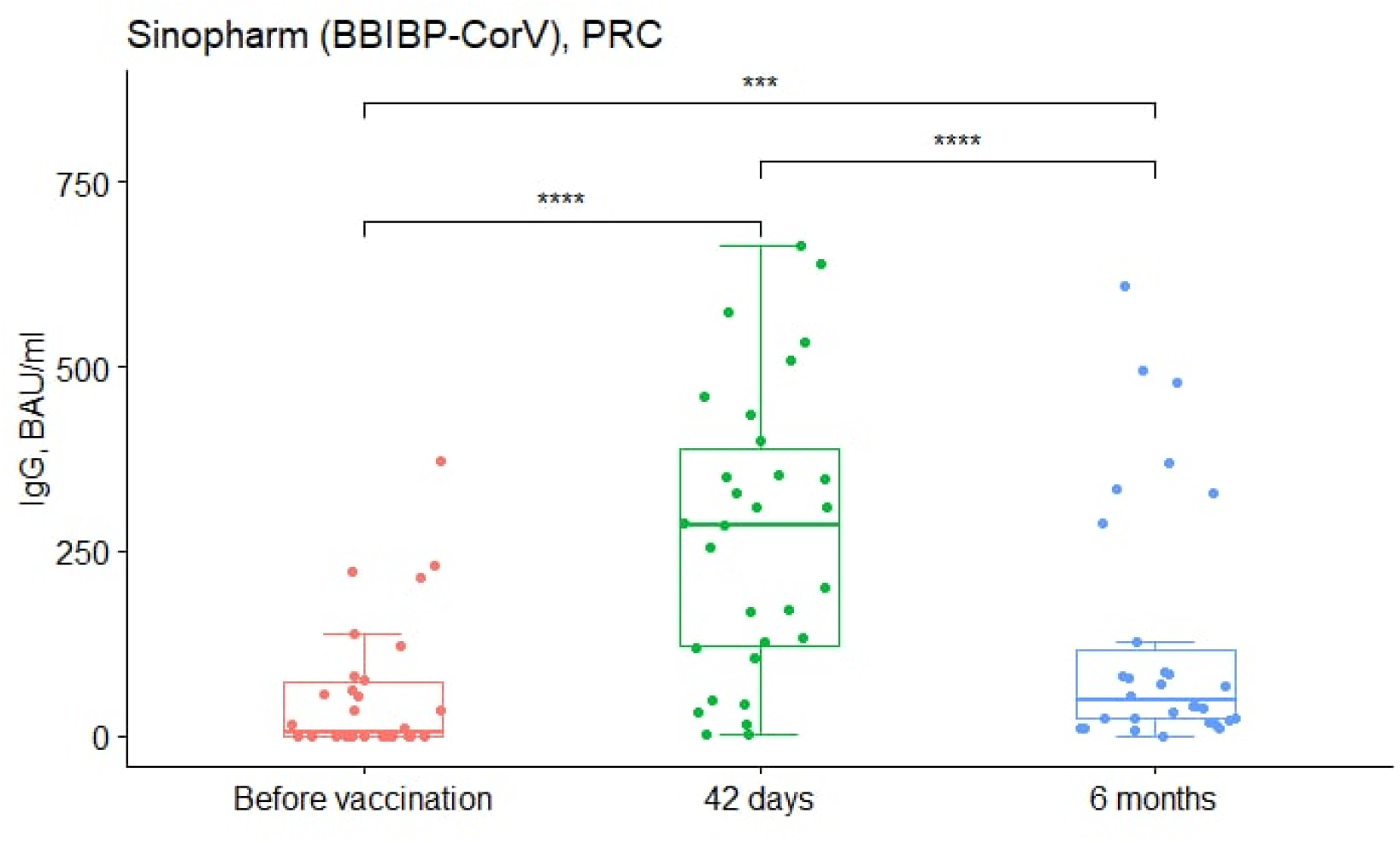
Dynamics of Ig G appearance in response to Sinopharm (BBIBP-CorV) vaccine (“before vaccination” - Ig G value (BAU/ml) before vaccination; “42 days” - Ig G value(BAU/ml) 42 days after the first dose; “6 months” - Ig G value (BAU/ml) 6 months after thefirst vaccine dose).

**Fig. 3.**
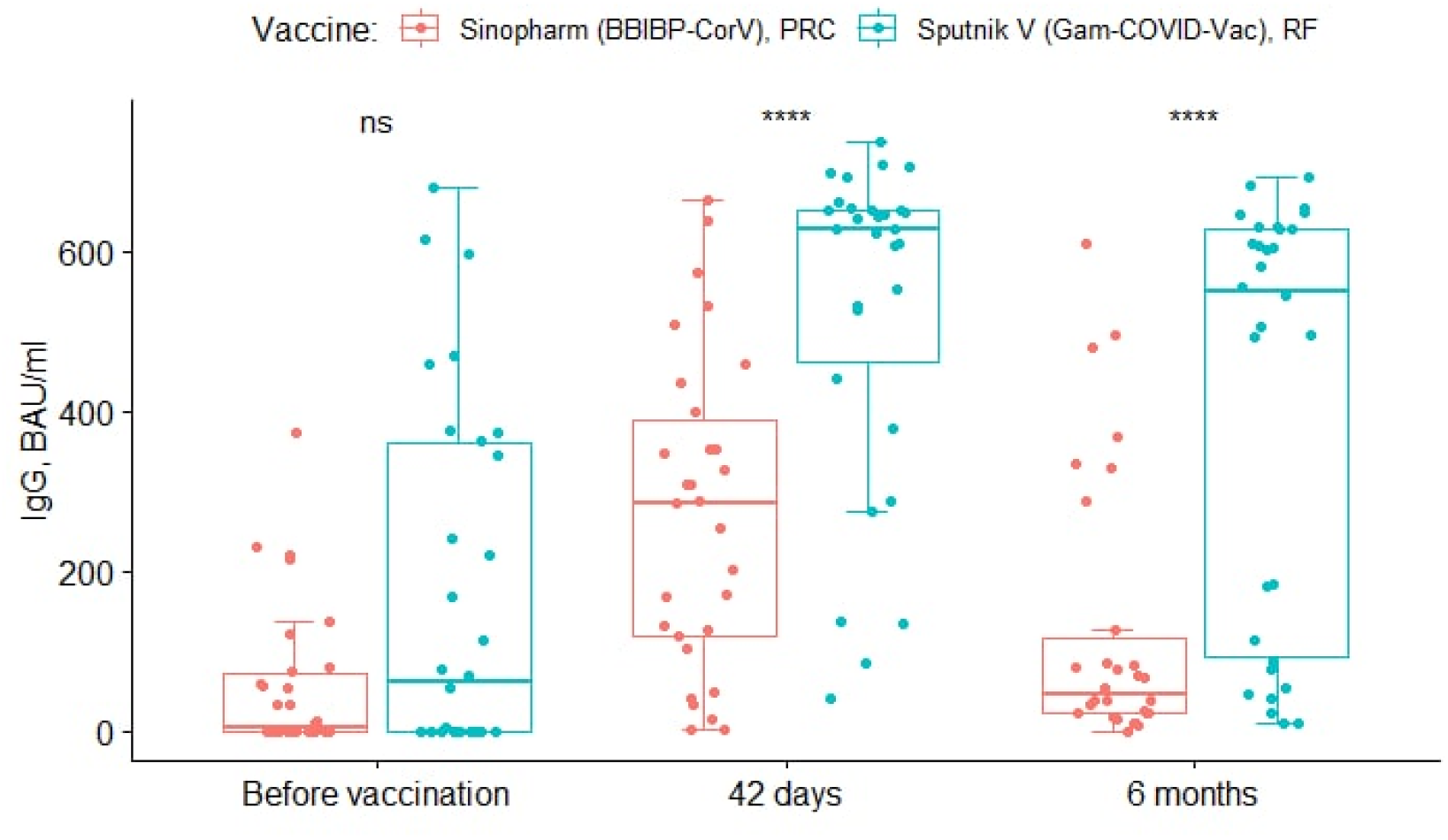
Comparison of the dynamics of Ig G levels in response to the administration ofSputnik V (Gam-COVID-Vac) and Sinopharm (BBIBP-CorV) vaccine (“Before vaccination” - Ig G value (BAU/ml) before vaccination; “42 days” - Ig G value (BAU/ml) 42 days after the first dose; “6 months” - Ig G value (BAU/ml) 6 months after the first vaccine dose).

As shown in Figure 1, quantitative IgG levels at the three measurement points have significant differences (Friedman chi-squared = 47.267, df = 2, p-value < 0,001). IgG levels 42 days after the first vaccine dose were significantly higher compared to the “Before vaccination” point (p<0,001), 6 months after the first dose of Sputnik V (Gam-COVID-Vac) vaccine the IgG level has a significant decrease (p < 0,001), however, compared to the “before vaccination” point, the IgG level remains at a higher level (p < 0,001).

As shown in Figure 2, the quantitative IgG values at the three study stages are significantly different (Friedman chi-squared = 44.891, df = 2, p-value < 0,001). Levels of IgG at 42 days after the first dose of vaccine were significantly higher than prior to vaccination (p < 0,001). At 6 months after the first dose, there was a significant decrease in IgG values (p < 0,001), but they remained at a higher level compared to the “before vaccination” “ point (p < 0,001).

At the pre-vaccination point there is no significant difference between the Sputnik V(Gam-COVID-Vac) and Sinopharm (BBIBP-CorV) vaccines (p=0.12). At 42 days after the administration of both vaccines, the antibody levels increase, but the quantitative IgG value for the Sputnik V (Gam-COVID-Vac) vaccine is significantly higher (p < 0,001). This trend was also observed 6 months after the first dose of both vaccines (p < 0,001).

According to a questionnaire survey answers of study participants, 51.67% of participants (31 people) did not have COVID-19 before vaccination. Accordingly, 48.33% of participants (29 people) had had the infection prior to vaccination. When comparing the groups characteristics on the basis of prior infection with SARS-CoV-2, no significant difference was found (p=0.071).

As the factor of previous SARS-CoV-2 infection may influence the outcome of the study, it was also included in the statistical analysis.

Figure 4 shows that in the Sputnik V (Gam-COVID-Vac) vaccine group, the largest group of individuals at the 42-day point had IgG levels above 500 BAU/ml, whereas most Sinopharm (BBIBP-CorV)-vaccinated patients had levels ≤500 BAU/ml (X-squared=19.644; df=2; p < 0,001). At the 6-month point, this trend continued: in the Sputnik V (Gam-COVID-Vac) vaccine group most individuals had IgG levels above 500 BAU/ml, while most patients vaccinated with Sinopharm vaccine had gG level below the 150 BAU/ml(X-squared=20.747; df = 2; p < 0,001). The groups were also compared taking into account the serological status of the study participants.

**Fig. 4.**
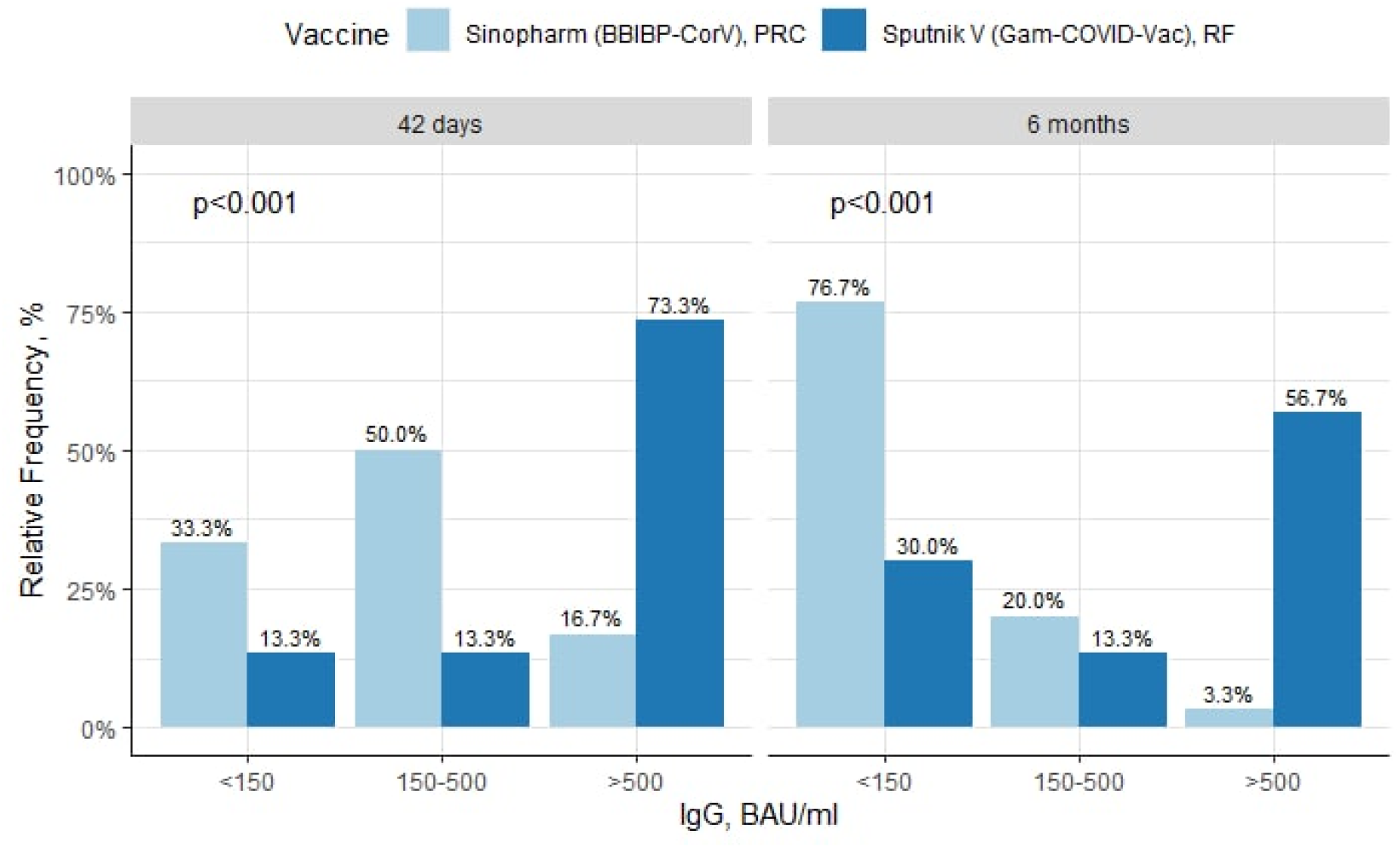
Comparison of the groups (vaccinated with Sputnik V (Gam-COVID-Vac) and Sinopharm (BBIBP-CorV)) by IgG levels (proportion reaching <150 BAU/ml, 150-500 BAU/ml, >500 BAU/ml) at study sites: “42 days” - 42 days after the first dose, “6 months” – 6 months after the first vaccine dose.

Figure 5 shows differences in IgG levels to the SARS-CoV-2 protein between those vaccinated with the Sputnik V (Gam-COVID-Vac) and Sinopharm (BBIBP-CorV) vaccines among both those who had not had COVID-19 and those who had had the infection before vaccination. The highest antibody levels were observed 42 days after vaccination in both the seronegative (p=0.006) and seropositive groups (p < 0,001), while 6 months after vaccination the IgG value declined among those who had not previously been ill (p=0.003) and those who had had COVID-19 before vaccination (p < 0,001).

**Fig. 5.**
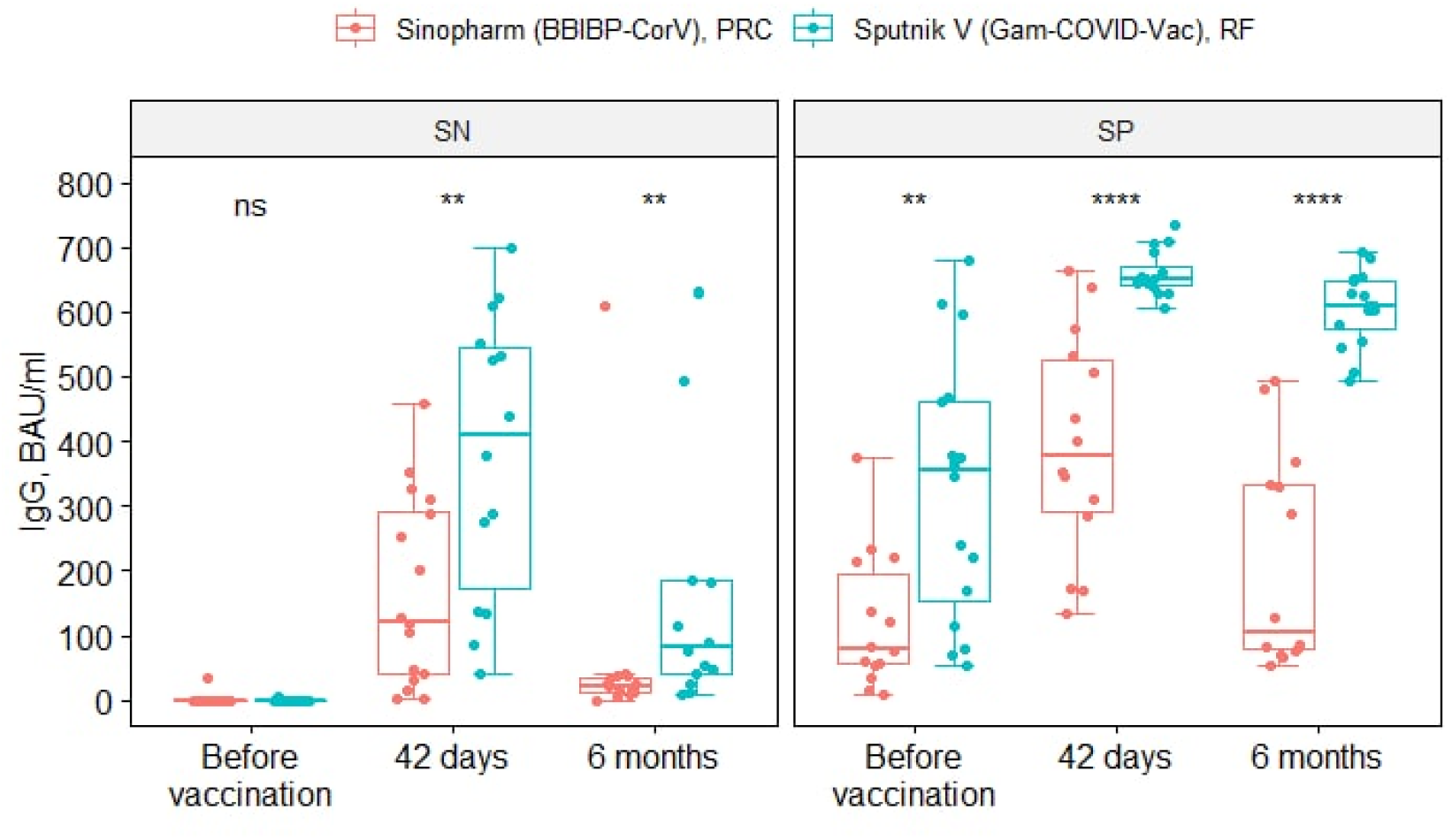
Comparison of Sputnik V (Gam-COVID-Vac) and Sinopharm (BBIBP-CorV) groups based on serological status before vaccination (“SN”-seronegative individuals, “SP”-seropositive participants; “Before vaccination” pre-vaccination IgG level, “42 days”-IgG level 42 days after the first dose of one of the vaccines, “6 months later”-IgG level 6 months after administration of one of the vaccines).

**Figure 6.**
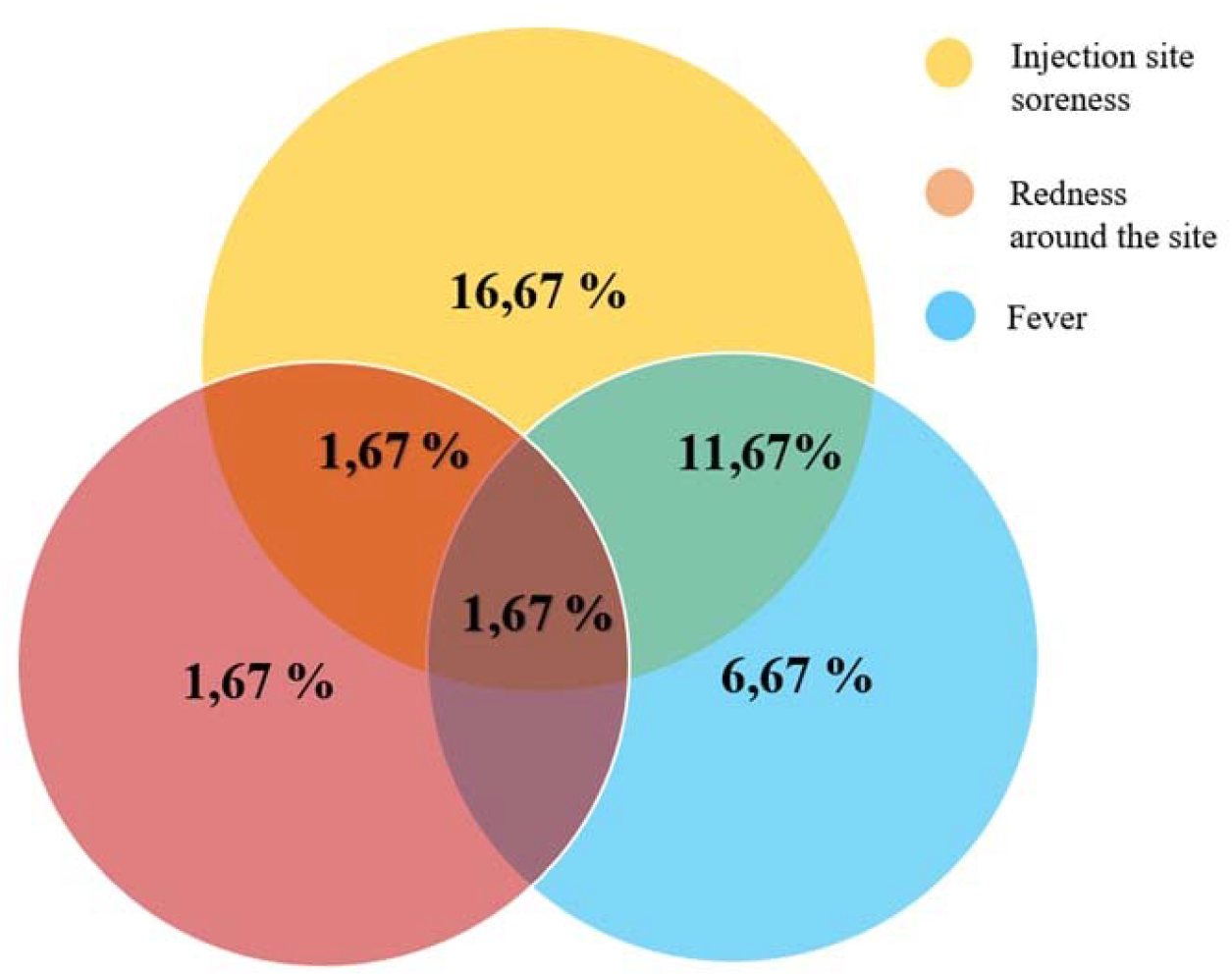
The main post-vaccination reactions (and their combination) noted in the study participants: soreness and redness at the injection site, increased body temperature.

There are a number of publications suggesting that those with “hybrid immunity” are most protected against COVID-19. Figure 5 also shows this trend. However, the “hybrid immunity” generated by the Sputnik V vaccine has greater strength and duration (p < 0,001).

The reactogenicity of Sputnik V (Gam-COVID-Vac) and Sinopharm (BBIBP-CorV)vaccines is worth considering separately, as this is a frequent topic of discussion. The incidence of post-vaccine reactions in this study was 45% (27 individuals). All of the observed reactions were mild to moderate in severity. The most frequent ones were: injection site soreness (16.67% of 10 people) and redness around the site of injection (5% - 3 people),fever (6.67% of 4 people), and a combination of these reactions (Fig. 8). Participants also reported weakness (1.67% - 1 person), headache (1.67% - 1 person), and swelling of the injection site (1.67% - 1 person).

Post-vaccination reactions were observed in both vaccinated groups (p=0.119). However, significant statistical differences were found only for the increase in body temperature: in the group vaccinated with Sputnik V (Gam-COVID-Vac) this reaction was more frequent (p=0.006).

## Discussion

Having evaluated both vaccines currently available to the population of Belarus, it can be concluded that the Sputnik V (Gam-COVID-Vac) vaccine is more immunogenic, but the Sinopharm (BBIBP-CorV) vaccine is less reactogenic. For a better understanding of the properties of the above mentioned vaccines, a comparative evaluation should be carried out.

Sinopharm (BBIBP-CorV) vaccine is an inactivated vaccine, which is the most traditional type of vaccine. The virus is grown in Vero cells, then purified and inactivated with β-propiolactone - this deprives the virus of its ability to reproduce, but retains its antigenic structure. The immune response of T cells is usually weak. Inactivated viruses enter host cells via endocytosis and stimulate helper T cells via the MHC II pathway, resulting in an activation of the humoral immune response. Inactivated vaccines do not normally stimulate cytotoxic T cells through the MHC-I pathway to any significant extent, so a cell-mediated immune response is not formed [9]. Inactivated vaccines require several doses and/or addition of adjuvant to achieve sufficient efficacy [10]. The adverse reactions described in the instructions are, in most cases, mild to moderate in severity, which resolve within a few days after vaccination. The most frequent reactions (≥1/10): headache, soreness at the injection site. Less frequent, but quite common (≥1/100 to <1/10): fever, muscle and joint pain, fatigue, cough, shortness of breath, nausea, diarrhoea, skin itching. The immunogenicty of the completed course of vaccination, as declared by the manufacturer, is 79.34%. Advantages include: 1) safety; 2) stability during transport and storage; 3) low cost. Disadvantages include: 1) low immunogenicity; 2) short-lived immune response [11].

Vaccine Sputnik V (Gam-COVID-Vac) vaccine is based on non-replicating viral vectors (adenoviruses 5 and 26) encoding the S-protein gene. Adenoviral vectors (Ad-vectors) have several features that make them ideal candidates: a high immunogenic profile and the ability, using a smaller amount of antigen, to induce both cellular and humoral immune response; Ad-vectors do not integrate into the host genome [12]. To create a vector, the viral E1 and/or E3 genes that enable replication are deleted and replaced with the transgene of interest (derived from the Wuhan-Hu-1 SARS-CoV-2 sequence) cloned together with a tissue plasminogen activator [13]. The efficacy of vector vaccines can be significantly affected by previous immunity. Vectors of a rare serotype (human adenoviruses Ad26, Ad5) are used for this purpose [12]. Advantages include: 1) high immunogenicity (without adding adjuvant); 2) stability (vectors used have a protein envelope that protects the genetic material inside it, hence the vaccine can be stored at +2 - +8 □ for at least 6 months; 3) possibility of large-scale production (Ad vectors can be grown in bioreactors of 20 l volume, yielding vaccine doses sufficient for 15,000 patients). Disadvantages include: 1) reactogenicity; 2) potential neutralization of vaccine vector by acquired immunity antibodies; 3) cost [14, 15]. The main adverse reactions that occur during the first days after administration of the Sputnik V (Gam-COVID-Vac) vaccine are mainly mild to moderate in severity, lasting about 3 days. The most frequent symptoms are: flu-like syndrome (chills, fever, arthralgia, myalgia, headache), soreness, hyperemia and swelling at the injection site. Less common are dyspepsia, nausea, nasal congestion, rhinorrhoea, sore and stuffy throat, enlargement of regional lymph nodes, and possible allergic reactions. The immunological efficacy of two doses of the vaccine, as declared by the manufacturer, is more than 91% (16).

Therefore, there is still a lack of international publications comparing the immunological efficacy and reactogenicity of Sputnik V (Gam-COVID-Vac) and Sinopharm (BBIBP-CorV) vaccines with each other. A retrospective observational study conducted in Mongolia found the immunogenicity of two doses of Sputnik V (Gam-COVID-Vac) vaccine to be significantly higher than the immunogenicity of two doses of Sinopharm vaccine. No reactogenicity study was conducted in this study (17). A retrospective observational study conducted in Hungary investigated the efficacy of Sputnik V (Gam-COVID-Vac), Sinopharm (BBIBP-CorV) et al. vaccines against SARS-CoV-2 infection (confirmed by PCR test) and death from COVID-19 among vaccinated individuals. The results of this study indicate that the Sputnik V (Gam-COVID-Vac) vaccine was 85.7% effective against SARS-CoV-2 infection and 95.4-100% effective against COVID-19 deaths (depending on age cohort), and the Sinopharm (BBIBP-CorV) vaccine was 68.7% effective against SARS-CoV-2 infection and 67.3-100% effective against COVID-19 deaths (depending on age cohort) [18].

Only few international publications reflect the results of a study of the reactogenicity of Sputnik V (Gam-COVID-Vac) and Sinopharm (BBIBP-CorV) vaccines. A study based on a survey of vaccinees conducted in Iran found that reactions in response to Sputnik V (Gam-COVID-Vac) vaccine were longer (up to 3 days) than reactions in response to Sinopharm vaccine (a few hours). The frequency of post-vaccination reactions also varied, with at least one reaction occurring in 93.2% of Sputnik V (Gam-COVID-Vac) vaccine users and 87.3% of Sinopharm (BBIBP-CorV) vaccine users. The most frequent reactions were pain at the injection site, headache and malaise. Also in the study there were cases of thrombosis and conditions related to clotting disorders: 7 cases after administration of Sputnik V (Gam-COVID-Vac) vaccine and 3 cases after administration of Sinopharm (BBIBP-CorV) vaccine. However, a subsequent association between vaccination and conditions related to clotting disorders has not been found for both vaccines, and this issue requires further investigation [19]. Another study, based on a survey of vaccinees vaccinated with Sputnik V (Gam-COVID-Vac), Sinopharm et al. in Iran, aimed to investigate cutaneous reactions in response to the administration of Sputnik V (Gam-COVID-Vac) and Sinopharm vaccines. Skin manifestations in the vaccinated subjects were transient. The most common reactions were pain at the injection site (79.9% of Sputnik V (Gam-COVID-Vac) vaccine users, 60.6% of Sinopharm vaccine users), tightening of the injection site (13.4% of Sputnik V (Gam-COVID-Vac) vaccine users, 10.3% of Sinopharm (BBIBP-CorV) vaccine users), reddening of the skin at the injection site (12.8% of Sputnik V (Gam-COVID-Vac) vaccine users, 6.9% of Sinopharm (BBIBP-CorV) vaccine users) [20].

Thus, the approaches to the study conducted at Gomel State Medical University do not contradict international studies and the study is conducted in a similar way, which in our opinion allows a comparative assessment of postvaccination reactions and immunogenicity of Sputnik V and Sinopharm vaccines.

The study has a number of limitations: sample size (60 people), age of participants (the study represents individuals whose age ranges from 29 to 73 years).

## Conclusions

According to the results of the study, the Sputnik V (Gam-COVID-Vac) vaccine compared to Sinopharm (BBIBP-CorV), has higher immunogenicity rates at both points: after 42 days (for Sputnik V (Gam-COVID-Vac) vaccine: Me=627.5 (461.7-650.8); for Sinopharm (BBIBP-CorV) vaccine: Me=286.0 (120.3-387.9); p=<0,001) and 6 months (for Sputnik V(Gam-COVID-Vac) vaccine: Me=549.4 (94.36-626.8); for Sinopharm (BBIBP-CorV) vaccine: Me=46.74 (22.23-116.4); p<0,001) after first dose administration. This trend was seen among both those previously infected with SARS-CoV-2 and those without a history of COVID-19.

The study confirmed that antibodies developed as a result of prior infection with SARS-CoV-2 combine with post-vaccination antibodies to form what is known as “hybrid immunity”. It creates higher levels of antibodies (for Sputnik V (Gam-COVID-Vac) (42 days) vaccine: Me=650.4 (642.2-669.4); for Sinopharm (BBIBP-CorV) (42 days) vaccine: Me=376.5 (290.9-526.4); p<0,001), which persist for a longer time (for Sputnik V (Gam-COVID-Vac) (6 mo) Me=608.7 (574.6-647.1); for Sinopharm (BBIBP-CorV) (6 months) vaccine Me=106.3 (78.21-332.4); p<0,001).

The main reactions after vaccination were fever, redness and soreness at the injection site. The reactogenicity of both vaccines was relatively similar (p=0,119), a fever after vaccination was more common among those vaccinated with the Sputnik V (Gam-COVID-Vac) vaccine (10 individuals or 33,33% vs. 3.33% of vaccinated with Sinopharm BBIBP-CorV, p=0.006).

Authors report no relevant conflict of interest regarding this study.

## Data Availability

All data produced in the present study are available upon reasonable request to the authors

## Acknowledgments

Authors would like to thank Alexey Zyatkov, Alexander Shaforost, Nadezhda Golubykh, Elena Lipskaya, Dmitry Redko, and Vera Shandyba for data collection and technical assistance.

